# A novel MRI-based three-dimensional model of stomach volume, surface area and geometry in response to gastric filling and emptying

**DOI:** 10.1101/2022.06.21.22276694

**Authors:** D Bertoli, EB Mark, D Liao, C Brock, JB Frøkjær, AM Drewes

**Affiliations:** Mech-Sense, Department of Gastroenterology and Hepatology, Aalborg University Hospital, Denmark; Department of Clinical Medicine, Aalborg University, Aalborg, Denmark; Mech-Sense, Department of Radiology, Aalborg University Hospital, Denmark

**Keywords:** Abdomen, Algorithms, Gastric emptying, Magnetic resonance imaging, Reproducibility

## Abstract

**Background:** Gastric motility and accommodation have a critical role in maintaining normal gastrointestinal homeostasis. Different modalities can be adopted to quantify those processes, i.e., scintigraphy to measure emptying time and intragastric Barostat for accommodation assessment. However, magnetic resonance imaging (MRI) can assess the same parameters non-invasively without ionizing radiation. Our study aimed to develop a detailed three-dimensional (3D) MRI model of the stomach to describe gastric volumes, surface areas, wall tension distribution, and inter-observer agreement.

**Methods:** Twelve healthy volunteers underwent an MRI protocol of six axial T2-weighted acquisitions. Each dataset was used to construct a 3D model of the stomach: Firstly, the volumes of the whole stomach, gastric liquid, and air were segmented. After landmark placing, a raw 3D model was generated from segmentation data. Subsequently, irregularities were removed, and the model was divided into compartments. Finally, surface area and 3D geometry parameters (inverse curvatures) were extracted. The inverse curvatures were used to as a proxy for wall tension distribution without measuring the intragastric pressure.

**Key Results:** The model was able to describe changes in volume and surface geometry for each compartment with a distinct pattern in response to filling and emptying. The surface tension was distributed non-homogeneously between compartments and showed dynamical changes at various time points.

**Conclusion & Inferences:** The presented model offers a detailed tool for evaluating gastric volumes, surface geometry, and wall tension in response to filling and emptying and will provide insights into gastric emptying and accommodation in diseases such as diabetic gastroparesis.

**KEY POINTS:**

- While MRI and ultrasound are getting progressively accepted as methods for evaluating gastric emptying and accommodation, they still do not provide essential insights into those processes.
- This study presents a three-dimensional stomach model able to report volume and surface data, and to describe the distribution of the gastric wall tension of gastric compartments by applying the Young-Laplace law.
- We observed that volumes and surface geometry showed distinct emptying patterns in each compartment and that the wall geometry distributed non-homogeneously in the stomach, showing different dynamical changes during the emptying phase. Our observations indicate the fundus as essential in the first phase of digestion, confirming its role as reservoir.
- The non-invasive model has the potential to give detailed information about the gastric volumes and surface geometry in response to filling and emptying, with the potential to understand the pathophysiology and improve treatment in patients with gastroparesis.

**GRAPHICAL ABSTRACT:** 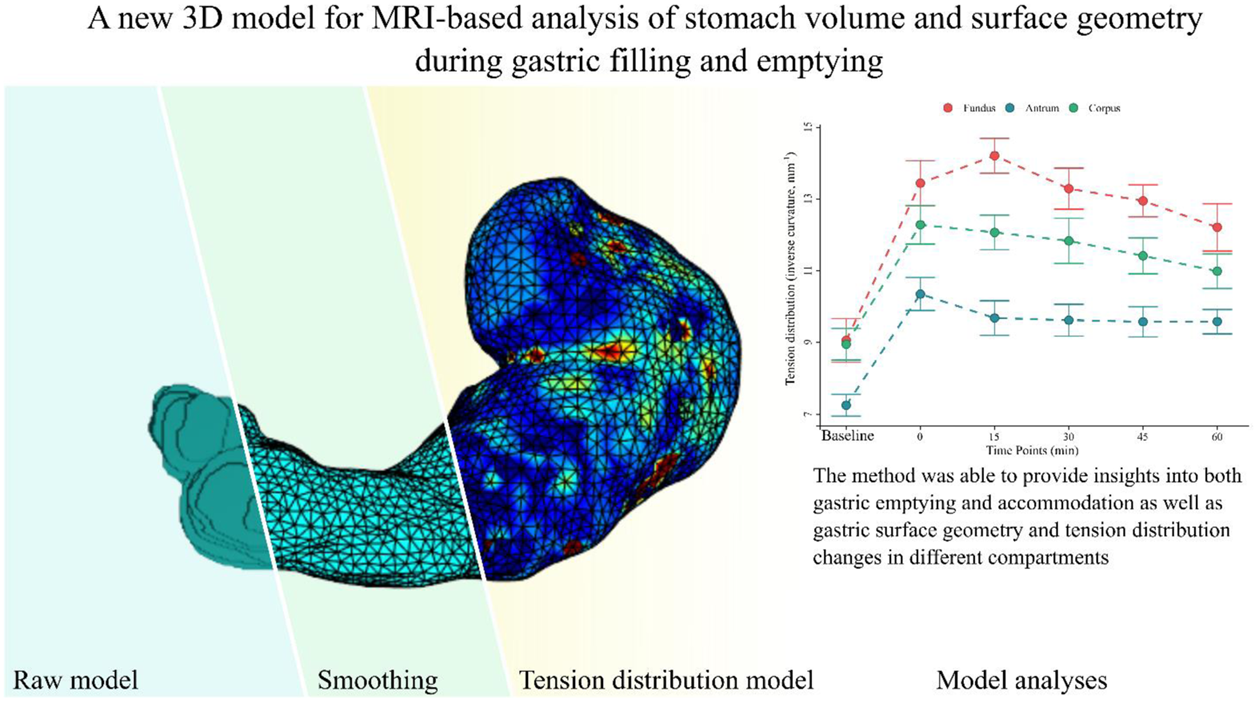

## INTRODUCTION

In modern practice, the interest in the function of the stomach has led to an increasing focus on the assessment of gastric emptying, motility, and accommodation, as these processes are closely associated with each other. Various examinations are commonly accepted such as: gamma scintigraphy,^1,2^ single-photon emission computed tomography,^3,4^ breath testing,^5,6^ intragastric Barostat,^7^ ultrasound,^8,9^ smartpill,^10^ and magnetic resonance imaging (MRI).^11,12^ Some of these methods are limited by invasiveness, the use of radioactive isotopes or by the inability to evaluate gastric emptying or anatomy. In contrast, others are time-consuming and offer only indirect measurements. Gamma scintigraphy is presently accepted as the gold standard in assessment of gastric emptying as it provides a reliable measure that does not suffer from confounding gastric secretions.^13^ Unfortunately, it does not offer a precise evaluation of gastric volumes, the contents’ intragastric distribution and wall geometry, resulting in a significant lack of insight into the mechanisms and pathophysiology behind diseases such as gastroparesis. Furthermore, its use is limited due to radiation exposure. The current standard for assessment of accommodation is the intragastric Barostat, an invasive technique not devoid of disadvantages and technical limitations.^7,14^

Because of these limitations, ultrasound and MRI are becoming progressively more attractive due to their ability to morphologically evaluate gastric emptying and motility, including intra-gastric volume distribution and geometry,^15–17^ without exposing the patient to unnecessary ionizing radiations or invasive procedures.^18^ However, ultrasound-based evaluation methods are prone to be observer-dependent and unable to visualize gastric air.^19^

MRIs, on the other hand, can visualize the entire stomach in one scan session providing three-dimensional (3D) morphological and functional information through repeated scans. While MRI examinations of the stomach and of the gastric function have been of interest in recent literature, the existing methods still have methodological limitations. For example, the intragastric pressure cannot be directly quantified and, therefore, the absolute wall tension value cannot be measured. Tension data are of utmost importance in the clinical application of these methods, as the gastric wall tension determines the perception of gastric distention.^20–22^ We hypothesized that an MRI 3D model would be able to provide essential insights into gastric geometry, motility, and accommodation. Furthermore, we hypothesized that through the application of the Young-Laplace law on the collected geometric data, the model would be able to describe the distribution of the wall tension across different compartments. Therefore, the primary aim of this study was to present a framework for a detailed analysis of gastric volumes, surface areas, and wall tension distribution in response to gastric filling and emptying using T2-weighted MR images before and after a liquid meal. The secondary aim was to apply the method to 12 healthy subjects, exploring the details of gastric filling and emptying processes. Finally, we aimed to measure the inter-observer agreement between two raters to further validate the observations on which our model was built.

## MATERIALS AND METHODS

### Study subject selection

Data were obtained from 12 healthy subjects, normal-weighted, without prior history of gastric disorders or other diseases affecting the gastrointestinal (GI) function. None of the subjects was treated with any medication that could affect the GI system. Informed consent was obtained per national and local institutions’ ethical standards. The study was approved by the North Denmark Region Committee on Health Research Ethics (N-20090008).

### Study design

Following a minimum of 6 hours of fasting (solids and liquids), a baseline scan (at t:-30 min) was obtained in a transversal plan with the subjects in supine position. Subsequently, subjects were asked to step outside the MRI scanner for 10 min and ingest a liquid meal of lightly heated 250 kcal tomato soup (500 ml “Karolines Køkken” (Arla Foods, Central Denmark Region, Denmark). Nutritional information per 100 g: fat 3g, carbohydrates 4.1g, proteins 1.1g. After ingesting the meal, the subjects were scanned at five additional time points (0, 15, 30, 45, and 60 minutes) with the same sequence as the baseline scan.

### MRI protocol

MRI scans were performed using a 1.5T General Electric model Discovery MR450 (General Electric Medical Systems, Milwaukee, WI, USA). Axial steady-state gradient echo (FIESTA) T2-weighted image series covering the entire stomach was obtained with an echo time (TE) = 1.5 ms and a repetition time (TR) = 3.5 ms, in-plane resolution = 0.7422 x 0.7422 mm, flip angle = 45°, slice thickness = 5 mm, 30-35 slices, and no image gap or overlap. Fat signal was not suppressed. Images were obtained in approximately 20 seconds under a single breath-hold to minimize respiratory artifacts.

As shown in previous similar studies,^23,24^ this sequence yielded a high signal of water and high contrast between gastric fluid content, comprehending meal and secretions, and gastric air as well as between the total gastric content and surrounding body tissues. No contrast-enhancing agents were deemed necessary to increase the accuracy of the measurement. An example of raw MRI images is shown in figure 1(A).

**Figure 1:**
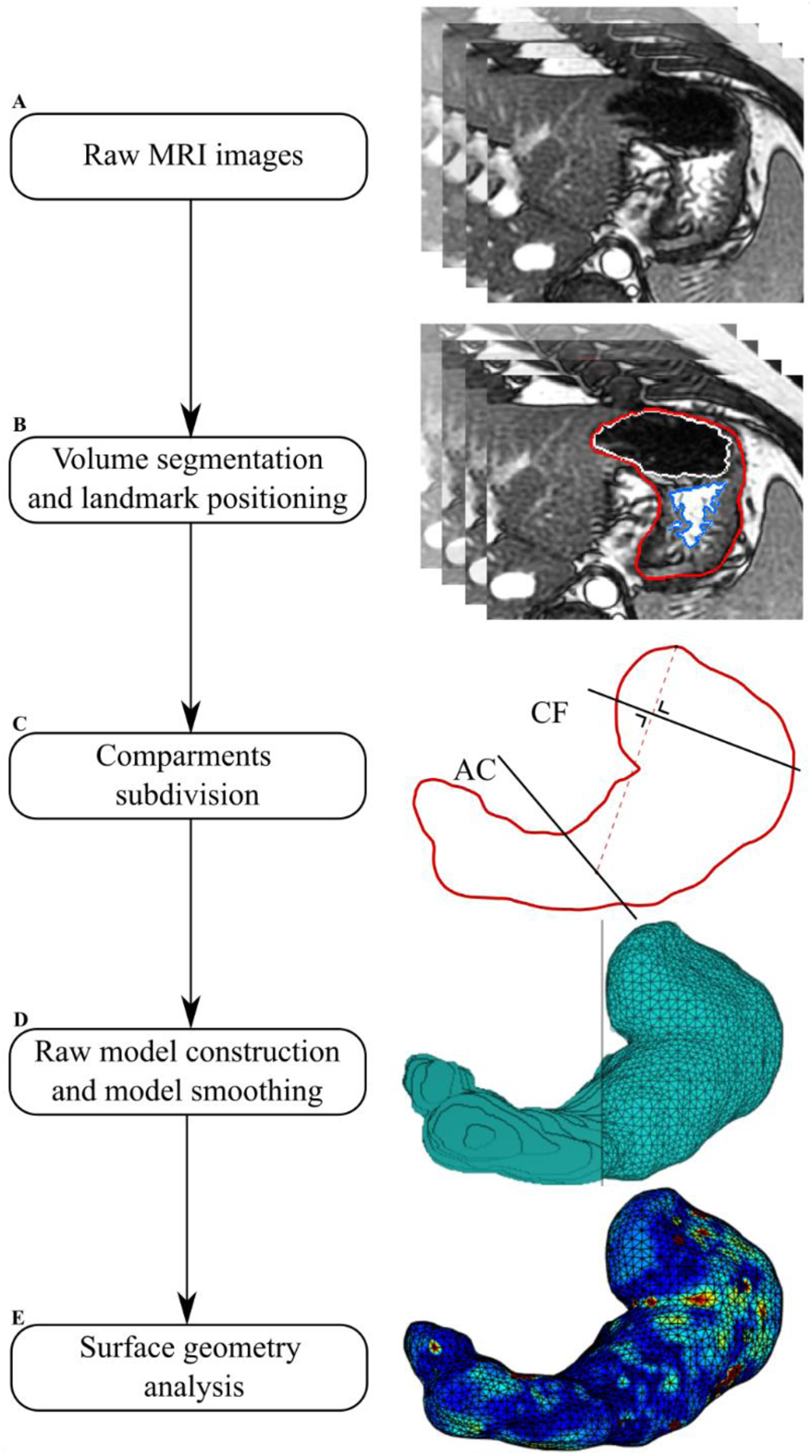
*Three-dimensional model development*. Firstly (A) axial T2-weighted images of the stomach were obtained. Then (B), the volumes of liquid gastric content (blue), air content (white), and total gastric content (red) were segmented. Landmarks at the central part of the cardia, angular incisure, and the topmost part of the stomach were placed in this phase (C). Secondly, a raw 3-dimensional model was generated from the segmented total gastric volume images (D). Irregularities were then removed, and the model was divided into compartments. Finally, inverse curvature data could be analyzed on the surface (E).

### Model preparation

#### Manual segmentation of gastric fluid content, gastric air, and total gastric volume

The segmentation of high-signal gastric fluid content, low-signal gastric air, and total gastric volume was manually performed using a segmentation platform implemented in MATLAB (v.R2018b, MathWorks, Natick, MA, USA). An example of the segmentation steps is shown in figure 1(B). The total gastric volume was defined by the outer border of the stomach, including the stomach wall. The gastric fluid content volume was defined as the high-signal volume within the total gastric volume (including areas with low signal caused by residues in the stomach or signal artifacts), while the gastric air was defined as the low-signal volume within the total gastric volume. Segmentations were performed by two observers (EBM and CS). Consensus was reached before further analysis. Firstly, segmentation of gastric fluid content and gastric air was performed. Secondly, an approximation of the total gastric volume boundary was initially drawn automatically using the previously segmented volumes to avoid redundant work. Then the boundaries were manually edited to include the gastric wall and gastric areas not included in the other volumes. Segmentations of all three compartments took approximately 2 hours per subject’s dataset, comprehending all 6 scans.

#### Compartments and geometry of the stomach

The stomach volume and surface models were divided into antrum, corpus, and fundus. The antrum was obtained by dividing the stomach with a plane (Antrum-Corpus plane, AC-plane) passing the angular incisure and perpendicular to the less curvature in the coronal plane, see figure 1(C). Fundus and corpus were divided by a plane (Corpus-Fundus plane, CF-plane) passing the central point of the cardia and perpendicular to the line between the topmost point of the stomach and the middle point of the AC-plane line in the coronal plane, see figure 1(C).^25,26^ The angular incisures used to divide the stomach into antrum, corpus and fundus were defined and independently confirmed by two observers (LD and EBM), and if there was any disagreement, consensus was reached before further analysis.

#### Three-dimensional volume and surface modelling of the stomach

3D raw stomach volume models were generated from the segmented total gastric volume data, see figure 1(D). The volume of the total stomach model was calculated in mL by multiplying the number of image pixels, pixel size, and slice thickness.

Based on the volume of the 3D stomach model, a first raw surface was computed using the isosurface function in MATLAB with an isovalue of 0. The point cloud obtained from the isosurface was down-sampled using a box grid filter in a box size of 5 mm and then presented as the surface of the stomach, represented with triangular facets, see figure 1(D). In order to remove the high curvature variations while avoiding shrinkage of the original surface, a modified non-shrinking Gaussian smoothing method was used to remove the irregularities of the reconstructed stomach surfaces due to the discretization of the images.^27^ The surface area of the entire stomach was then calculated in cm^2^ by adding up the areas of all triangular facets.^28^ The same method was previously applied, e.g., in a work of Liao et al.^29^ At this step of our pipeline, geometric data were extracted from the model, and inverse curvature data was calculated. For details of surface computations, see Appendix 1: *surface smoothing*. The processing of geometric data will be discussed in the following sections.

### Data processing

#### Gastric emptying half-times

The change of gastric liquid, air, and total gastric volume during the emptying phase was curve-fitted to a linear-exponential model (LinExp),^30^ allowing to quantify the gastric emptying half-times as:

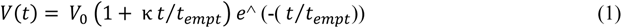

This model can handle an initial volume increase (due to gastric secretions) using the coefficient kappa (κ) and the subsequent volume decrease described by the *t_empt_* coefficient.^24^ The *V*_0_ coefficient describes the start volume (t:0 min).

This calculation was performed with the online tool’ apps.menne-biomed.de/gastempt/’. This tool has been previously utilized in other studies.^31^ In this work, the fitting method *nlme population fit* was utilized.

#### Wall tension distribution

In this work, the gastric wall tension was not directly measured or estimated but through the following principles and equations it was possible to describe how the tension was distributed in different compartments (called *Wall tension distribution* in this work). In medicine, the Young-Laplace equation (2) has been used to describe multiple phenomena like the formation of diverticula or the expansion of abdominal aorta aneurysms^32^:

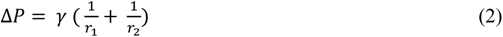

where ΔP is defined as Δ*P* = *P_i_* − *P*_o,_ where *P*_o_ is the intra-abdominal pressure and *P_i_* is the intragastric pressure; assuming isotropic tensions, *γ* is the tension on the gastric wall, and r_1_ and r_2_ are the principal radii of curvature.

In differential geometry, the principal curvatures at a given point on a surface are the maximum and minimum values of all the possible curvatures obtained through the section of the surface by planes tangent to the surface containing the normal vector 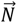. These two curvatures are denoted k_1_ and k_2_ and can be expressed as:

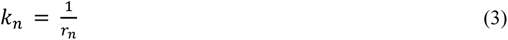

Integrating (3) in (2):

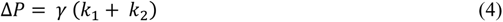

Resolving (4) for the ratio *γ* / Δ*P*, (5) is obtained:

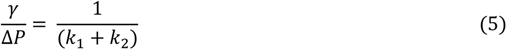

Introducing in (5) the mean curvature, an extrinsic measure of curvature defined in fluid mechanics as:

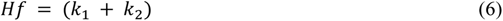

We obtain that:

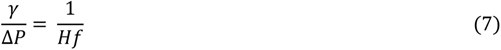

Furthermore, assuming P_o_ and P_i_ (and therefore Δ*P*) to be constant (*c*) within each timepoint, the tension would only be proportional to the inverse mean curvature, allowing us to describe the wall tension distribution in different gastric compartments within the same timepoint, given only geometric data:

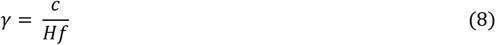

and therefore:

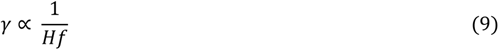

To report this distribution, inverse curvature data were normalized to antral values at the same timepoint, as we expected the volumes to be accommodated mainly in the proximal stomach.^26^ As the constant *c* is of unknown value, this method does not allow us to estimate absolute wall tension values but allows us to evaluate its distribution on the gastric surface. For a visual representation of the forces involved, see figure 2. *k*_1_ and *k*_2_ (expressed as mm^−1^) were calculated from the 3D surface model of the stomach using the algorithm presented by Hartmann et al.^28^ See Appendix 1: *principal curvatures computation*. These analyses were at last applied to the previously generated 3D model. See figure 3 and figure 1(E).

**Figure 2:**
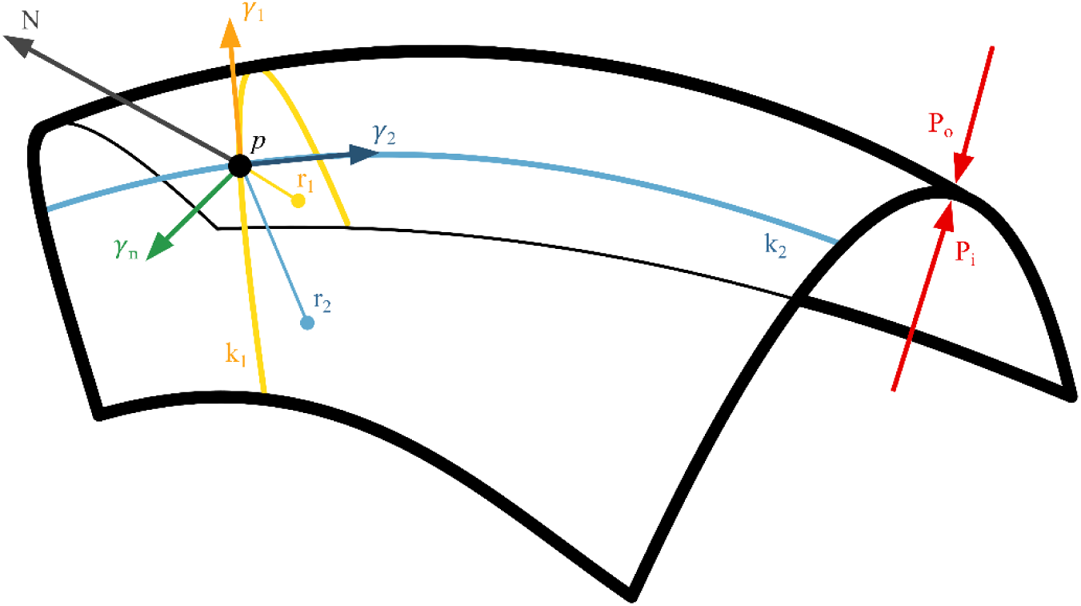
*Schematic representation of the physical forces involved in the accommodation process*. P_o_: abdominal pressure, P*_i_*: intragastric pressure, k_1_ and k_2_ are the principal curvatures with r_1_ and r_2_ their relative radii, N is the normal vector perpendicular to the surface, *γ* is the tension on the gastric wall.

**Figure 3:**
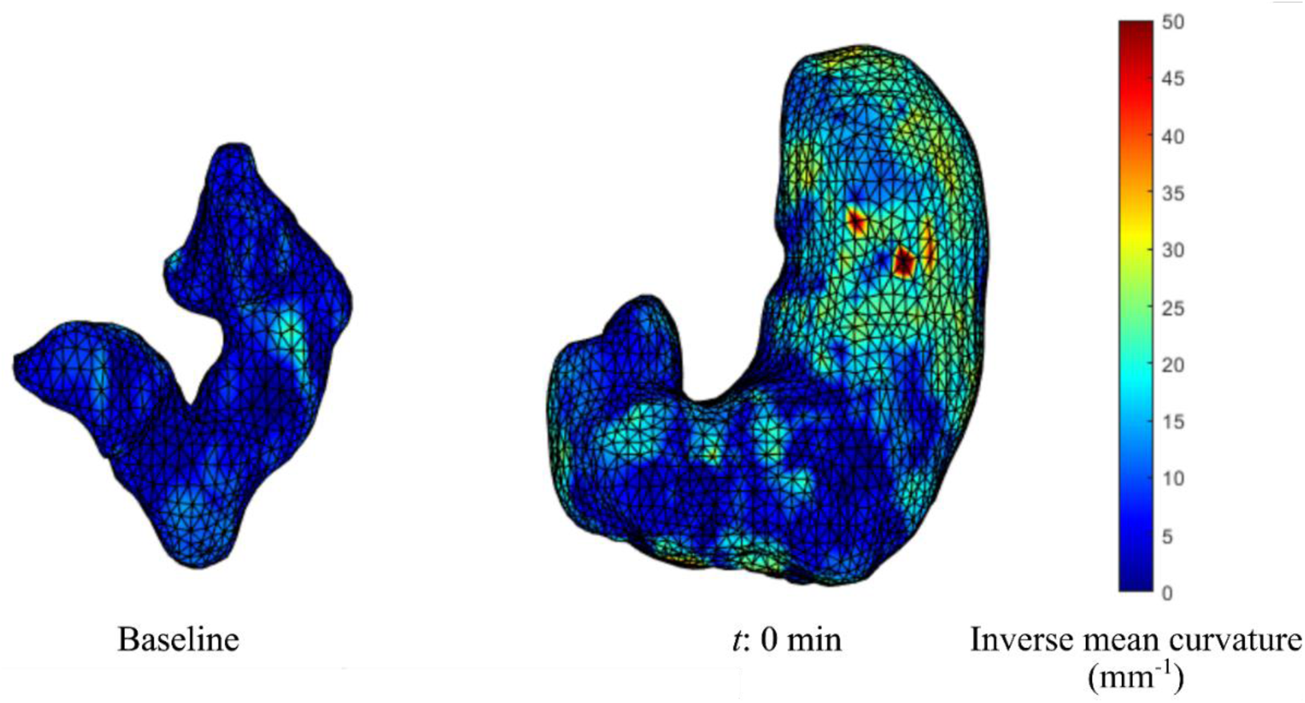
*Tension distribution*. The image shows an example of an empty stomach at baseline, characterized by lower wall tensions (blue color due to low inverse mean curvature values), compared to an image of a full stomach at t:0 min, characterized by higher tensions, particularly in the fundus (yellow/red colors due to high inverse mean curvature values).

Another important assumption in our model is that the stomach was represented as a thin-walled membrane. This assumption is based on the premise that to properly apply the Laplace’s law on a non-infinitesimal-walled structure, the following requirement should be met:

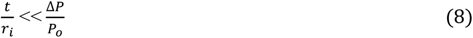

where *t* is the wall thickness, *r_i_* the internal radius, and *P*_o_ the intra-abdominal pressure.

Studies reported mean intra-abdominal pressures *P*_o_ of 1.7 (1.2) cmH_2_0 and mean intra-gastric pressures *P_i_* of 2.9 (1.7) cmH_2_O, resulting in 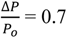 in the pre-swallow resting state.^33,34^ Resulting in:

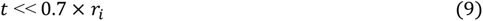

This requirement was met in our dataset. Wall thickness data were not reported as outside the scope of this work.

Applying these equations to the geometric data collected allow to describe the distribution of the wall tension in different compartments without measuring or estimating the intragastric pressure.

### Data normalization

Volume and surface area data were normalized to their total values to better convey the compartment dynamics during the emptying phase. The inverse curvature data were normalized to antral values at the same timepoint (see *Wall tension distribution* above).

### Inter-observer agreement

As the initial part of this framework is heavily observer-dependent, inter-observer differences in the total gastric volume were explored through Dice similarity coefficients. For validation, intraclass correlation coefficient (ICC) assessments between the two raters (EBM and CS) were performed on the extracted geometric data (inverse curvatures) on which our model was built. Data were obtained by two different raters (EBM and CS). No limit of agreement was established a priori.

### Statistical analysis

Data were analyzed in R (R Core Team, 2021, R Foundation for Statistical Computing, Vienna, Austria) with *rstatix* package (R package version 0.7.0), and figures were produced using the package ggplot2.^35^

After identifying and removing extreme outliers (defined as values above the third quartile + 3 interquartile ranges or below the first quartile – 3 interquartile ranges), volume, surface area, and inverse curvature data were inspected for normality through Q-Q plots and Shapiro-Wilk tests. Correlation tests were then performed with two-way repeated-measure analysis of variances (ANOVAs, *time* and *compartment* were used as independent variables, dependent variable: volume, surface areas, or inverse curvature, both non-normalized and normalized), tested for sphericity with Mauchly’s test, and corrected with Greenhouse-Geisser and Huynh-Feldt corrections. In the case of positive correlation, posthoc analyses were performed with pairwise t-tests adjusted with sequential Bonferroni tests (Bonferroni-Holm). A *p*-value < 0.05 was considered statistically significant. Data were reported as mean (SD). ICC estimates and their 95% confidence intervals were calculated using R packages *blandr* and *psych*.^36,37^ The model ICC1 (single-measure, absolute-agreement, two-way mixed effect) was utilized. Data was not transformed for analysis and were visually reported through a Bland-Altman plot.

## RESULTS

All the recruited subjects fully complied with the study protocol, and a complete dataset was obtained from all twelve subjects (7 males), mean age 27.8 years (range 23-34 years). All the subjects well tolerated the meal.

### Gastric volumes

Volumes of gastric fluid content, gastric air, and the total gastric volume can be seen in figure 4(A).

**Figure 4:**
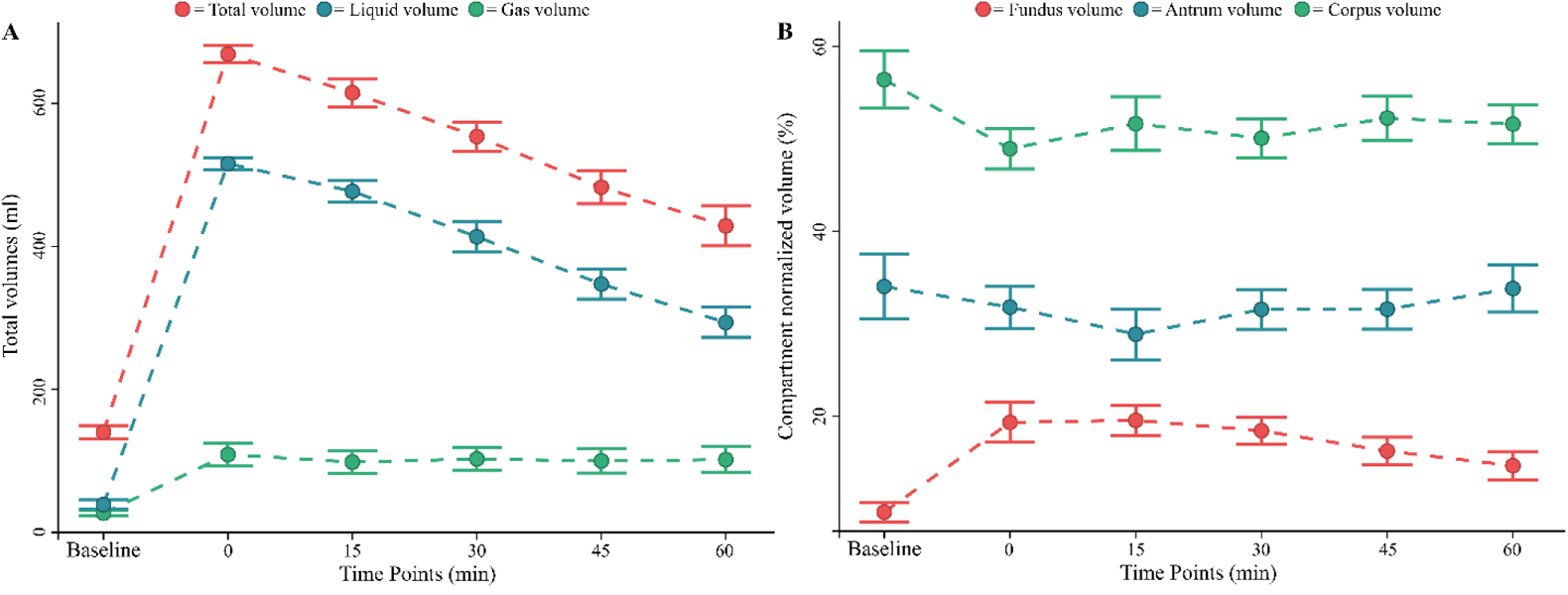
*Gastric volume data*. A: total gastric volume (red), total liquid volume (blue), and total gas volume (green) are shown. B: Normalized fundus volume (red), normalized corpus volume (green), and normalized antrum volume (blue) are shown. All data are reported as mean ± 1 standard error (error bars).

The total gastric volume increased from baseline values 140.3±31.9ml to 669.1±41.4ml at t:0 min (*p* < .001) and decreased to 428.7±96.8ml at t:60 min (*p* < .001), remaining however over baseline values (*p* < .001). The initial increase in volume was mostly due to the ingested meal: the liquid volume increased from 38.7±23.2ml at baseline to 515.8±29.9ml at t:0 min (*p* < .001). The liquid volume showed a similar decreasing pattern with a minimal volume of 293.6±73.9ml at t:60 min, never reaching baseline values (*p* < .001).

The gastric gas volume, after a limited initial increase from baseline values of 26.8±13.7ml to 108.7±54.9ml at t:0 min (*p* < .001), remained constant and over baseline values throughout the examination (*p* < .001), with a final volume of 101.7±62.9ml at t:60 min.

All compartments’ total volumes (comprehending gastric wall, liquid, and air content) showed a similar dynamic with an initial increase (all *p* <.001) from baseline values to t:0 min and a successive decrease to t:60 min (all *p* <.05). No compartment volume returned to baseline values at t:60 min (all *p* <.05).

Data showed a difference at baseline between the volumes of each compartment with a fundus volume of 13.4±6.0ml, corpus volume of 80.4±26.7ml, and antrum volume of 46.5±18.2ml (all *p* <.001). This observation was confirmed by a two-way repeated ANOVA that showed an influence of both time and compartment localization on the compartment volumes (all *p* < .001). Normalized data showed that fundus volumes increased from 9.6±3.6% at baseline to 19.3±7.5% at t:0 min and never returned to baseline values throughout the examination, with a volume of 14.6±5.3% at t:60 min. No change from baseline values of corpus (56.4±10.7%) or antrum (64.0±12.0%) was observed (all *p* > .05). The different dynamics can be observed in figure 4(B).

The complete dataset can be found in Appendix 2, Table A1.

### Assessment of emptying half-times

The emptying half-times of the liquid volume showed a mean value of 69.3 (SD 15.2) minutes. The emptying half-times of the total gastric volume showed a mean value of 86.0 (SD 18.0) minutes.

### Gastric surfaces

The total gastric surface area increased from 220.4±27.2cm^2^ at baseline to 536.4±25.0cm^2^ at t:0 min (*p* < .001) and decreased to 413.4±54.2cm^2^ at t:60 min. See figure 5(A).

**Figure 5:**
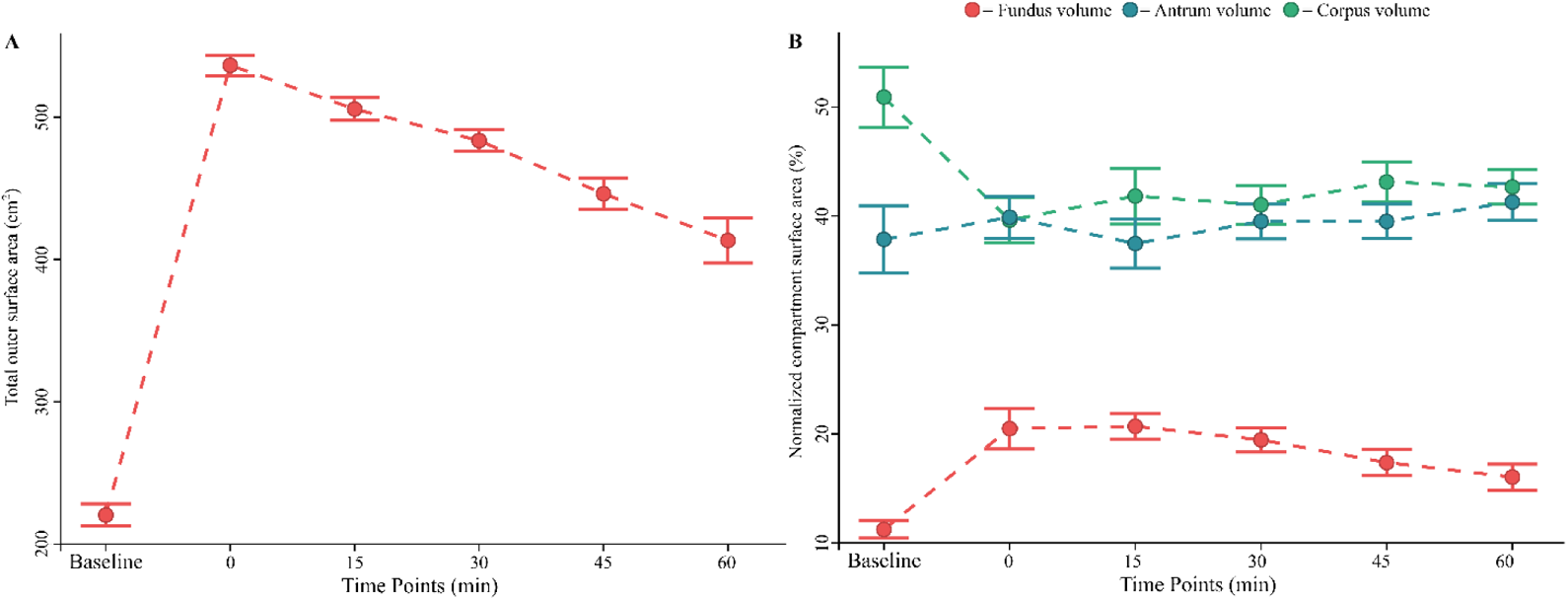
*Gastric surface area data*. A: Total gastric surface area is shown. B: Normalized fundus surface area (red), normalized corpus surface area (green), and normalized antrum surface area (blue) are shown. All data are reported as mean ± 1 standard error (error bars).

All compartmental surface areas showed a similar dynamic with a starting increase from baseline to t:0 min (all *p* < .001) and a successive decrease toward t:60 min (all *p* < .05).

Fundus surface area was smaller than the other compartmental areas (all *p* < .001) with a baseline value of 24.5±5.7cm^2^ for fundus, 111.8±22.6cm^2^ for corpus, and 84.2±30.8cm^2^ for antrum. This observation was confirmed by a two-way repeated ANOVA that showed an influence of both time and compartment localization on the compartment surface (all *p* < .001). Normalized data showed that the fundic surface area increased from 11.2±2.8% at baseline to 20.7±4.1% at t:15 min (*p* = .001), never returning to baseline values throughout the examination. This increment was mirrored by an initial decrease of corpus surface area from 50.9±9.6% at baseline to 41.8±8.9% at t:15 min (*p* < .001). Corpus surface area never returned to baseline throughout the study. No change was observed in antrum surface area, which remained around baseline (37.9±10.7%) during the examination. The different surface area dynamics can be seen in figure 5(B).

The complete dataset can be found in Appendix 2, Table A2.

### Gastric wall tension distribution

The wall tension distribution was different between compartments (*p* < .001), with mean inverse curvature values in the fundus being higher than those in the corpus (*p* < .001) and corpus values being higher than those in the antrum (*p* < .001) at every timepoint. The dynamical changes in the tension distribution can be seen in figure 6(A) for non-normalized data and figure 6(B) for normalized data. Data showed that the corpus tension was 26±26% higher than the antral tension (*p* = .017). At t:0 min, the tension in the fundus increased to values 34±34% higher than those in the antrum (*p* = .018), while corpus tension values still were 21±24% higher than antrum ones (*p* = .026). Fundus tension continued to increase to t:15 min to 51±32% of the antral tension (*p* < .001). From t:15 min, the tension distributed toward the antrum with values in fundus and corpus steadily diminishing throughout the examination to t:60min.

**Figure 6:**
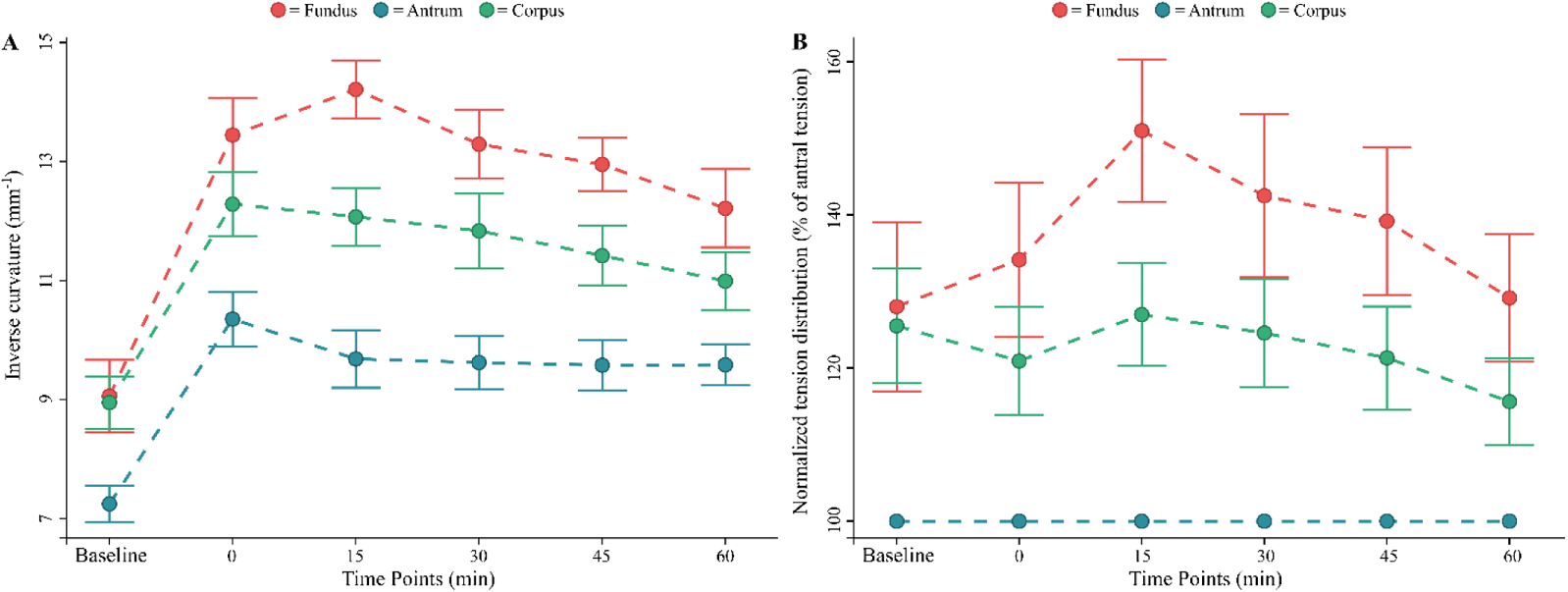
*Inverse curvature data*. A: the non-normalized values of fundus (red), corpus (green), and antrum mean inverse curvature value (blue) are shown. B: fundus (red) and corpus (green) inverse curvatures are shown normalized to antral (blue) values at the same timepoint.

The complete dataset can be found in Appendix 2, Table A3.

### Inter-observer agreement

Data showed a median Dice similarity coefficient for total gastric volume of 0.957 (IQR = 0.021) between the two raters.

Dice similarity coefficients for liquid and gas can be seen in figure 7(A). Median values ranged from baseline values: 0.846 (0.047) for total gastric volume, 0.851 (0.101) for liquid volume, and 0.828 (0.020) for gas volume, to 0.965 (0.009) for total gastric volume, 0.975 (0.013) for liquid volume, and 0.902 (0.050) for gas volume at t:0 min.

**Figure 7:**
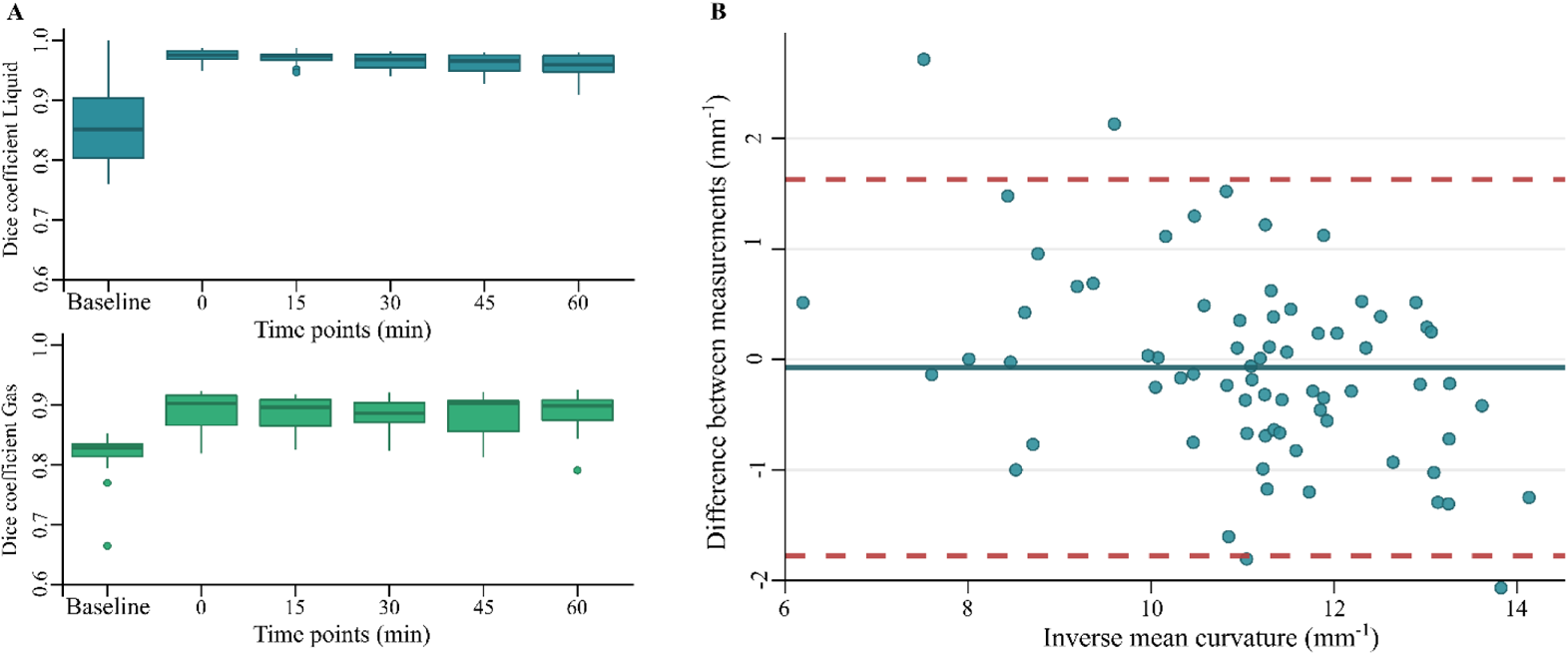
*Dice similarity coefficient and intraclass correlation coefficient data*. A: Dice similarity coefficients for the liquid (blue) and gas (green) volumes are shown. B: Bland-Altman plot for the correlation agreement between the two inverse curvature datasets across every timepoint. Blue dots: observations, blue line: bias, red dashed lines: 95% limits of agreement.

ICC validation results can be seen in figure 7(B). The estimated bias was −0.07mm^−1^ (−0.28, 0.13), with lower limit of agreement: −1.78mm^−1^ (−2.12, −1.43) and upper limit:1.63mm^−1^ (1.28, 1.98). The correlation coefficient was 0.87 (lower bound 0.79, upper bound 0.91, *p* < .005).

## DISCUSSION

The present study presents a 3D MRI-based model for a detailed geometric assessment of gastric compartment volumes, surface areas, and wall tension distribution using MRI. The most important findings study were: 1) The model was able to describe changes in volume, surface areas, and wall tension distribution in fundus, corpus, and antrum; 2) The changes in the observed gastric variables in response to gastric filling and emptying showed a distinct pattern for each compartment; 3) The wall tension distributed non-homogeneously between different compartments and showed different dynamical changes during the emptying phase. Furthermore, during gastric filling, we observed that the most significant changes in volume and surface area occurred in the fundus, underlining its role as a reservoir of undigested food. The geometric data on which our model was built also gave us important insights into the dynamics of the emptying process, as, during the first 15 minutes after meal intake, the normalized fundus tension increased, representing its predominant role in the first phase of the emptying process.

### Gastric volumes and emptying

As expected, the volume of the ingested liquids played a major role in the observed total gastric volume increase, while the gas volume was only accessory and generally constant throughout the examination. Furthermore, we observed that the fundus played a major role in accommodating newly ingested meals, as it was the only compartment to increase its normalized volume and surface area values during the first 15 min. The observed fasting volumes were in line with the literature,^38,39^ as were the dynamics of the emptying process.^40^ The volume and dynamics of total gas content were also in line with findings observed in the literature.^26^ With a careful approximation (due to differences in the compartment subdivision of the stomach), it appears that the antral volumes at the start of the filling phase were also in line with the literature.^26^

MRI gastric emptying studies have reported emptying half-times between 7 and 330 minutes in healthy subjects for liquid and mixed meals.^41^ Our half-time measurements of 69.3 min for liquid and 86.0 min for the total volume comply with these observations. Direct comparison with previous data is difficult due to differences between study protocols, such as different amounts and kinds of meals,^23^ MRI sequences,^25^ or different sample sizes or demographics.^30^ Moreover, some studies positioned the patients in a right lateral decubitus, which permits an easier outflow through the antrum,^42^ and it is not always stated which positions were used between each scan.^43^ However, the emptying phase dynamic of the fundus, corpus, and antrum in the present study showed a good agreement with a recent study done by Banerjee et al.^26^

### Gastric surface areas analysis

While no standard reference values of surface areas can be found in the literature, the dynamic of our observations is in line with other recent studies.^44^ In our work, the fundus appears to sustain the most relevant area increase after meal intake, as it was the only compartment to show an increase in its normalized surface area from baseline to t:0 min.

Previously, we have done geometric surface analyses on 3D imaging data from the stomach,^45^ gallbladder,^46^ and rectum in humans to investigate the geometry and mechanical change of the models during distension.^47^ In our previous surface modelling analysis, we re-sliced the 3D volume model along the central line of the model to generate the surface, which was very time-consuming.^47^ The method presented in this study has a superior time efficiency compared to the previous modelling analysis as it implemented a surface smoothing which could be performed straightforwardly from the volume model reconstructed from the MR images.

### Gastric tension distribution

In our observations, the gastric tension was distributed unevenly across different compartments, with the highest inverse curvature values in the fundus and the lowest in the antrum. Furthermore, we observed that during the earliest phase of digestion (15 min), the fundus reached tension values up to 51% higher than those observed in the antrum. These observations concords with the reported changes in volume and surface areas elicited in the fundus by the meal, indicating the fundus as the most influential compartment in the first phase of digestion, confirming its role as reservoir. Our data showed wide variations, as observable by relatively high standard deviations. As data were normally distributed and the inter-observer variability was excellent (see *Inter-observer agreement* below), these differences can be imputed to significant inter-subject variability, which is also often observed in other tracts of the GI system.^10,48,49^

### Inter-observer agreement

Our analysis showed an excellent agreement in total gastric volumes between the two observers, defined by an overall Dice similarity coefficient of 0.938. Our observed coefficient is significantly higher than 0.7, defined in the literature as a good overlap.^50^ ICC analysis also showed good reliability, defined as an ICC value between 0.75 and 0.9.^51^

### Potential clinical implications

Implementation of analytical tools like the currently described image acquisition and analysis framework in the clinical practice will require optimization of the analysis time, including optimized segmentation and landmark annotation. A more automatic and standardized method will rapidly assess gastric function, emptying, and other motility indexes to evaluate and diagnose motility disturbances. These disturbances range from delayed gastric emptying and gastroparesis to abnormally rapid transit, usually referred to as “dumping syndrome”.^52,53^ Furthermore, it will give access to the assessment of the gastric accommodation process, which plays an essential role in functional dyspepsia symptoms or in gastroesophageal reflux.^15^ These assessments are vital in clinical practice and pharmacological studies.^54^ Also, more automatic and standardized methods could be helpful in future research studies, giving them access to fast and straightforward 3D gastric models that could lead to a more robust understanding of the motility and the relative interferences of diseases or potential treatment approaches (drugs, neuromodulation, gastric pacing, and others).

### Study limitations

In the present study, the subjects were placed in supine position after meal intake in the following image acquisitions. This position displaces the gastric content away from the antrum, potentially slowing gastric emptying.^42^ A possible improvement could be obtained by oblique positioning in Fowler’s position (semi- or low-) as that would offer a more realistic setting for the gastric environment.^55^ However, in the majority of similar studies, the study subjects were also placed in supine position as this is easier with standard MRI equipment.^41^

An unavoidable issue in the segmentation of gastric volumes is the lack of a precise anatomical boundary between the duodenum and pylorus, making it challenging to outline identical segmented compartments of the stomach during all six scans. Custom boundaries were adopted as previously described, but their overall impact on measurements is still unclear.^56^

Due to the lack of a direct measurement of the intragastric pressure, estimating exact absolute wall tension values was not possible in this study. This issue could be overcome by measuring the intragastric pressure before and after food intake.^57^ Furthermore, more insights into the stomach’s motility could be assessed through cine-imaging or through thinner slices, which could potentially be implemented in our framework.

### Conclusions

As per our aims, the analysis framework presented in this study offered a tool for evaluating gastric volumes, surface areas, and a dynamic estimation of wall tension based on MRI, feasible with only minor preparation. The method was able to provide insights into both gastric accommodation and emptying. Compared to existing gastric emptying studies, our model is improved by estimating the gastric wall tension distribution changes in different stomach compartments and holds promise for future clinical implementations and research.

## DISCLOSURE

No competing interest declared.

## AUTHOR CONTRIBUTIONS

Davide Bertoli, Esben Bolvig Mark, Jens Brøndum Frøkjær, Donghua Liao, and Asbjørn Mohr Drewes designed the research; Davide Bertoli, Esben Bolvig Mark, and Donghua Liao were involved in data collection; Asbjørn Mohr Drewes, Davide Bertoli, Christina Brock, and Jens Brøndum Frøkjær directed the data interpretation; Davide Bertoli drafted and wrote the manuscript; All the authors revised the final manuscript.

## Data Availability

All data produced in the present study are available upon reasonable request to the authors

## ACKNOWLEDGMENTS

We thank Kenneth Krogh Jensen (Aalborg University Hospital) for the assistance in the data collection and the Radiology Research Unit for providing MRI scanner facilities.

We thank Christoffer Svinth for the assistance in the data collection.

