## Appendix 1 for "A novel MRI-based three-dimensional model of stomach volume, surface area and geometry in response to gastric filling and emptying"

***Surface smoothing***

The reconstructed surfaces had some irregularities due to the discretization of the images, which were removed using a modified non-shrinking Gaussian smoothing method. The relation between the position of the vertices before and after N iteration can be expressed as:

$X^{N}={(\left( I-\mu K \right)\left( I-\lambda K \right))}^{N}X$	(A1)

where *N* was the number of iterations, $\lambda$ and $\mu$ are two scale factors, *I* is the *n_v_* $\times$*n_v_* identity matrix, *K = I - W*, *W* is the weight matrix, and *n_v_* is the number of the neighborhood of a vertex. In this study, the scale factors and the iteration number ranged from 20 to 50. They were selected according to the criterion that the relative error between the surface area calculated from the smoothed model and the raw model must be lower than 10%.

***Principal Curvatures Computation***

The stomach has a complex 3-D geometry but since the surface is smooth and continuous, it can be approximated locally by a quadric surface function as:

F(*x,y,z*) = x^2^ + a_1_y^2^ + a_2_z^2^ + a_3_x + a_4_y + a_5_z + a_6_ = 0	(A2)

Where a_i_ (*i* = 1, 2, …, 6) are constants. For each vertex, its 3rd-folds neighborhood (including vertexes and faces) was first defined. Secondly, the vertex normal was calculated by averaging the faces normals of the neighborhood. The surface area of each face was then weighted by dividing it by the summarized face areas of the neighborhood. The previously defined vertex normal was then utilized to transpose the vertex and its 3rd-folds neighborhood vertexes within a local coordinate system (o-xyz). This system was defined with the vertex as the origin point and the vertex normal as the z-direction of the local coordinate system. The constants a*_i_* (*i* = 1, 2, …, 6) were obtained by least-squares fitting of the surface function (A2) to the vertex and its 3^rd^-folds neighborhood points in the local coordinate system. Hence, the principal curvatures of the vertex can be calculated from the coefficient of the first fundamental form (E, F and G) and the second fundamental form (L, M and N) of the differential geometry as:

$K_{G}= k_{1}k_{2}= \frac{LN- M^{2}}{EG- F^{2}}$(A3)

$K_{M}=\left( k_{1}+ k_{2} \right)= \frac{NE-2MF+LG}{EG- F^{2}}$ (A4)

Where $k_{1}$ and $k_{2}$ are the principal curvatures, $K_{G}$ and the $K_{M}$ are *Gaussian* and *Mean* curvatures, respectively. K_G_ is a particularly useful curvature parameter that indicates an elliptical surface (K_G_ > 0), a parabolic surface (K_G_ = 0) or a hyperbolic surface (K_G_ < 0). K_M_ is in inverse proportion to the surface tension according to the Laplace's Law, $\Delta P= \gamma\left( k_{1}+ k_{2} \right),$ where $\Delta P$ denotes the transmural pressure acting on the surface, $\gamma$ is the surface tension assumed constant in every direction.

Based on the calculation of fundamental forms of an implicit surface as presented by Hartmann:

*The coefficient of the first fundamental form (E, F and G) are*:

$E=1+\frac{F_{x}^{2}}{F_{z}^{2}}$ , $F=1+\frac{F_{x}F_{y}}{F_{z}^{2}}$ , and $G=1+\frac{F_{y}^{2}}{F_{z}^{2}}$	(A5)

Where F is the surface function in A2, F_x_, F_y,_ and F_z_ are the first-order partial derivatives of F.

*The coefficient of the second fundamental form (L, M and N) are*:

$L = \frac{1}{F_{z}^{2}\left| \nabla F \right|}\left| \begin{matrix} F_{xx} & F_{xz} & F_{x} \\ F_{zx} & F_{zz} & F_{z} \\ F_{x} & F_{z} & 0 \end{matrix} \right|, M= \frac{1}{F_{z}^{2}\left| \nabla F \right|}\left| \begin{matrix} F_{xy} & F_{yz} & F_{y} \\ F_{zx} & F_{zz} & F_{z} \\ F_{x} & F_{z} & 0 \end{matrix} \right|, N= \frac{1}{F_{z}^{2}\left| \nabla F \right|}\left| \begin{matrix} F_{yy} & F_{yz} & F_{y} \\ F_{zy} & F_{zz} & F_{z} \\ F_{y} & F_{z} & 0 \end{matrix} \right|$ (A6)

Where $\left| \nabla F \right|=\sqrt{F_{x}^{2}+F_{y}^{2}+F_{z}^{2}}$ and F_xx_, F_yx_ = F_xy_, F_yy_, F_yz_ = F_zy_, F_zz_, and F_xz_ = F_zx_ are the second-order partial derivatives of the surface function F in A2.
