## Appendix 2 for "A novel MRI-based three-dimensional model of stomach volume, surface area and geometry in response to gastric filling and emptying"

***Table A1***

| Time | Total gastric volume  (ml) | Total  liquid  volume  (ml) | Total  gas  volume  (ml) | Fundus  volume  (ml) | Corpus volume  (ml) | Antrum  volume  (ml) | Fundus normalized  volume  (%) | Corpus normalized  volume  (%) | Antrum normalized volume  (%) |
| --- | --- | --- | --- | --- | --- | --- | --- | --- | --- |
| Baseline | 140.3  (31.9) | 38.7  (23.2) | 26.8  (13.7) | 13.4  (6.0) | 80.4  (26.7) | 46.5  (18.1) | 9.6  (3.6) | 56.4  (10.7) | 34.0  (12.0) |
| 0 | 669.1  (41.4) | 515.8  (29.9) | 108.7  (54.9) | 130.1  (51.6) | 327.1  (51.4) | 110.1  (50.5) | 19.3  (7.5) | 48.9  (7.7) | 31.8  (7.9) |
| 15 | 614.9  (68.8) | 477.0  (53.6) | 98.2  (55.8) | 120.2  (36.6) | 315.7  (58.9) | 104.6  (64.4) | 19.5  (5.6) | 51.6  (10.1) | 28.8  (9.5) |
| 30 | 553.8  (70.6) | 413.5  (74.2) | 102.6  (56.4) | 101.8  (28.9) | 276.1  (49.0) | 93.8  (50.3) | 18.4  (5.0) | 50.1  (7.2) | 31.5  (7.4) |
| 45 | 482.8  (79.9) | 347.3  (72.2) | 99.8  (58.3) | 79.7  (32.9) | 250.4  (48.1) | 77.7  (44.0) | 16.2  (5.2) | 52.2  (8.3) | 31.6  (7.5) |
| 60 | 428.7  (96.8) | 293.6  (73.9) | 101.7  (62.9) | 64.2  (30.2) | 219.1  (45.0) | 66.8  (53.1) | 14.6  (5.3) | 51.6  (7.3) | 33.8  (8.8) |

Table A1. ***Gastric volume data***. The table shows total, compartmental, and normalized compartmental volumes. Data are reported as mean (SD).

***Table A2***

| Time | Total gastric  surface  (cm^2^) | Fundus  surface  (cm^2^) | Corpus  surface  (cm^2^) | Antrum  surface  (cm^2^) | Fundus  normalized  surface  (%) | Corpus  normalized  surface  (%) | Antrum  normalized  surface  (%) |
| --- | --- | --- | --- | --- | --- | --- | --- |
| Baseline | 220.4  (27.2) | 24.5  (5.7) | 111.8  (22.6) | 84.2  (30.8) | 11.2  (2.8) | 50.9  (9.6) | 37.9  (10.7) |
| 0 | 536.4  (25.0) | 110.1  (36.1) | 212.2  (36.7) | 214.1  (37.1) | 20.5  (6.5) | 39.6  (7.1) | 39.9  (6.6) |
| 15 | 505.7  (27.7) | 104.6  (21.4) | 210.6  (40.5) | 190.4  (44.2) | 20.7  (4.1) | 41.8  (8.9) | 37.5  (7.8) |
| 30 | 483.5  (26.3) | 93.8  (17.9) | 198.0  (30.0) | 191.7  (33.5) | 19.5  (3.9) | 41.0  (6.2) | 39.5  (5.6) |
| 45 | 446.2  (37.3) | 77.7  (21.0) | 191.7  (27.4) | 176.8  (31.4) | 17.4  (4.2) | 43.1  (6.4) | 39.5  (5.4) |
| 60 | 413.4  (54.2) | 66.8  (20.9) | 174.9  (21.5) | 171.7  (40.2) | 16.0  (4.2) | 42.7  (5.6) | 41.2  (5.9) |

Table A2. ***Gastric surface area data***. The table shows total, compartmental, and normalized compartmental surface areas. Data are reported as mean (SD).

***Table A3***

| Time | Total inverse curvature  (mm^-1^) | Fundus  inverse curvature (mm^-1^) | Corpus  inverse curvature (mm^-1^) | Antrum  inverse curvature (mm^-1^) | Fundus  normalized  inverse curvature (%) | Corpus  normalized  inverse curvature (%) | Antrum  normalized  inverse curvature (%) |
| --- | --- | --- | --- | --- | --- | --- | --- |
| Baseline | 8.6  (1.1) | 9.1  (2.1) | 9.0  (1.5) | 7.2  (1.1) | 28.0  (38.2) | 25.5  (25.9) | 1  (0) |
| 0 | 12.0  (0.8) | 13.4  (2.2) | 12.3  (1.9) | 10.4  (1.6) | 34.1  (34.7) | 20.9  (24.4) | 1  (0) |
| 15 | 11.9  (1.0) | 14.2  (1.7) | 12.1  (1.7) | 9.7  (1.7) | 51.0  (32.1) | 27.0  (23.2) | 1  (0) |
| 30 | 11.5  (1.2) | 13.3  (2.0) | 11.8  (2.2) | 9.6  (1.5) | 42.5  (36.9) | 24.6  (24.4) | 1  (0) |
| 45 | 11.2  (0.9) | 13.0  (1.5) | 11.4  (1.7) | 9.6  (1.5) | 39.2  (33.5) | 21.3  (23.3) | 1  (0) |
| 60 | 11.0  (0.9) | 12.2  (2.3) | 11.0  (1.7) | 9.6  (1.2) | 29.1  (28.8) | 15.6  (19.6) | 1  (0) |

Table A3. ***Gastric inverse curvature data***. The table shows total, compartmental, and normalized compartmental inverse curvature. Normalized data is reported as the increased percentage reported to antral data at the same timepoint.
